# Deep Learning Utilizing Suboptimal Spirometry Data to Improve Lung Function and Mortality Prediction in the UK Biobank

**DOI:** 10.1101/2023.04.28.23289178

**Authors:** Davin Hill, Max Torop, Aria Masoomi, Peter J. Castaldi, Edwin K. Silverman, Sandeep Bodduluri, Surya P. Bhatt, Taedong Yun, Cory Y. McLean, Farhad Hormozdiari, Jennifer Dy, Michael H. Cho, Brian D. Hobbs

## Abstract

**Background:** Spirometry measures lung function by selecting the best of multiple efforts meeting pre-specified quality control (QC), and reporting two key metrics: forced expiratory volume in 1 second (FEV_1_) and forced vital capacity (FVC). We hypothesize that discarded submaximal and QC-failing data meaningfully contribute to the prediction of airflow obstruction and all-cause mortality.

**Methods:** We evaluated volume-time spirometry data from the UK Biobank. We identified “best” spirometry efforts as those passing QC with the maximum FVC. “Discarded” efforts were either submaximal or failed QC. To create a combined representation of lung function we implemented a contrastive learning approach, **Spiro**gram-based **C**ontrastive **L**earning **F**ramework (Spiro-CLF), which utilized all recorded volume-time curves per participant and applied different transformations (e.g. flow-volume, flow-time). In a held-out 20% testing subset we applied the Spiro-CLF representation of a participant’s overall lung function to 1) binary predictions of FEV_1_/FVC < 0.7 and FEV_1_ Percent Predicted (FEV_1_PP) < 80%, indicative of airflow obstruction, and 2) Cox regression for all-cause mortality.

**Findings:** We included 940,705 volume-time curves from 352,684 UK Biobank participants with 2-3 spirometry efforts per individual (66.7% with 3 efforts) and at least one QC-passing spirometry effort. Of all spirometry efforts, 24.1% failed QC and 37.5% were submaximal. Spiro-CLF prediction of FEV_1_/FVC < 0.7 utilizing discarded spirometry efforts had an Area under the Receiver Operating Characteristics (AUROC) of 0.981 (0.863 for FEV_1_PP prediction). Incorporating discarded spirometry efforts in all-cause mortality prediction was associated with a concordance index (c-index) of 0.654, which exceeded the c-indices from FEV_1_ (0.590), FVC (0.559), or FEV_1_/FVC (0.599) from each participant’s single best effort.

**Interpretation:** A contrastive learning model using raw spirometry curves can accurately predict lung function using submaximal and QC-failing efforts. This model also has superior prediction of all-cause mortality compared to standard lung function measurements.

**Funding:** MHC is supported by NIH R01HL137927, R01HL135142, HL147148, and HL089856.

BDH is supported by NIH K08HL136928, U01 HL089856, and an Alpha-1 Foundation Research Grant.

DH is supported by NIH 2T32HL007427-41

EKS is supported by NIH R01 HL152728, R01 HL147148, U01 HL089856, R01 HL133135, P01 HL132825, and P01 HL114501.

PJC is supported by NIH R01HL124233 and R01HL147326.

SPB is supported by NIH R01HL151421 and UH3HL155806.

TY, FH, and CYM are employees of Google LLC

## 1 Introduction

The measurement of lung function via spirometry is one of the most common diagnostic tests used in clinical practice. Spirometry is required for diagnosis of COPD, and is a key diagnostic test used in assessing asthma, interstitial lung disease, neuromuscular disease, preoperative pulmonary risk, and monitoring for drug toxicity. While the number of spirometry tests performed is not known, spirometry at least annually is recommended for asthma and COPD, diseases that have a combined prevalence of over nearly 500 million people. Lung function is described using summary measurements from raw volume-time spirometry curves. Spirometry is performed by an individual first taking a maximum inhalation and then performing a maximum forced expiratory effort into a spirometry device. Spirometry measurements are often noisy with repeated efforts typically resulting in variable summary measures of lung function including the forced expiratory volume in 1 second (FEV_1_) and the forced vital capacity (FVC). Spirometry guidelines, as they can depend on effort, and in contrast to most other biomedical measurements, require repetition. In clinical practice, individuals are required to perform a minimum of two high-quality and reproducible (volume variability less than a specified threshold) spirometry efforts, and often perform three or more. The “best” FEV_1_ and FVC are then selected as the highest measured value for each measure across all high-quality and reproducible efforts. Spirometry quality control (QC) guidelines include factors such as minimal back-extrapolated volume (i.e. assuring the forced exhalation effort is maximal from the very start of the exhalation maneuver), sufficient amount of time performing the forced exhalation, and a sufficient volume plateau at the end of the forced exhalation effort. Any spirometry efforts that are not reproducible or do not meet these QC guidelines are discarded, since FEV_1_ and FVC are only extracted from the maximal efforts.

In addition to omitting submaximal and QC-rejected spirometry efforts, traditional spirometry measurements only summarize a few features of the spirometry curve, usually the FEV_1_ (the volume of forcibly exhaled air in the first second of an effort) and the FVC (the total forcibly exhaled volume), which may not capture a comprehensive representation of lung function. Alternative methods have been investigated using different aspects of the spirometry curve (Vandevoorde et al., 2008; Simon et al., 2010) to see if alternatives to the FEV_1_ and FVC are better associated with important clinical outcomes and disease progression. Machine learning and deep learning methods have been applied to raw spirometry curves for specific tasks, such as prediction of Chronic Obstructive Pulmonary Disease (COPD) (Bhattacharjee et al., 2022), COPD subtyping (Bodduluri et al., 2020), prediction of upper airway obstruction (Wang et al., 2022a), or acceptability criteria (Das et al., 2020; Wang et al., 2022b). In these applications, the models are trained exclusively on a single representative spirometry effort from each individual.

We hypothesize that the discarded submaximal and QC-rejected spirometry efforts contain useful information related to lung function and overall health outcomes, which can be captured using deep learning methods trained on the totality of the spirometry curves. While the information carried by discarded efforts may be degraded compared with the maximal effort, that information may still be relevant to understanding overall lung function. Further, the combined information from the discarded efforts may be sufficient to reconstruct the information from the maximal effort, which would reduce the number of efforts required during a clinical visit. Additionally, the discarded efforts also contain information related to the variance and reproducibility of the reported spirometry values. The magnitude of variance and the existence of failed efforts may be correlated to underlying lung function and similarly, to pulmonary disease. Thus, information mined from discarded spirometry efforts may provide a more comprehensive metric of an individual’s lung function.

Therefore, the goal of this project was to determine whether the information from these discarded efforts would provide comparable or even superior information on lung function and mortality. To do this, we first investigated the relationship between information captured from discarded spirometry efforts and the information classically reported from the maximal or “best” spirometry effort. We used deep learning methods on the raw spirograms from the UK Biobank to extract a vector representation of lung function for each individual.

In order to assess the quality of the deep-learning derived vector representation of lung function, we applied it in two different prediction applications: 1) incidence and severity of airflow obstruction and 2) mortality prediction. In each of these prediction tasks, the vector representation of lung function is used as a replacement for alternative spirometry metrics including the commonly used FEV_1_ and FVC.

Our results showed that including the discarded spirometry efforts in the vector representation of lung function led to improved prediction of lung function impairment and mortality prediction compared to both utilizing only maximal efforts and classic summary measures of lung function from spirometry.

## 2 Methods

### 2.1 Data Quality Control and Preprocessing

We extracted raw spirometry values from the UK Biobank field 3066. We used only spirometry data from the initial visit for each participant, thus excluding multiple visits from the same participant. Raw spirometry data were recorded as total exhaled lung volumes (mL) at 10 ms intervals over a median of 7.9 seconds and a range of 0.3 to 30.0 seconds (App. D Fig. 11). Each participant had between one and three spirometry efforts.

The following quality control filtering and preprocessing steps were applied to the spirometry samples. The spirometry QC was performed in line with the American Thoracic Society (ATS) and European Respiratory Society (ERS) guidelines using the procedure previously described by Shrine et al. (2019). QC steps are detailed in Table 2. For model validation purposes, we required all participants to have at least one QC-passing and reproducible effort. Spirometry efforts were flagged as passing or failing each step of the QC process (Fig. 1). After each step, participants with less than one passing effort were removed from the dataset.

**Table 1:** Characteristics of participants in the study.

| Characteristic | Training Set | Validation Set | Test Set |
| --- | --- | --- | --- |
| Participants, n | 211,603 | 70,534 | 70,535 |
| Spirometry Efforts, n | 564,509 | 188,105 | 188,091 |
| Participants with 3 Efforts, n (%) | 141,279 (66.8) | 47,037 (66.7) | 47,021 (66.7) |
| Age, yr, mean (SD) | 56.3 (8.1) | 56.3 (8.1) | 56.3 (8.1) |
| Sex, F, n (%) | 118,064 (55.8) | 39,528 (56.0) | 39,454 (55.9) |
| Ethnicity, Self-Reported |  |  |  |
| White, n (%) | 201,074 (95.3) | 66,968 (95.2) | 66,963 (95.2) |
| Black or Black British, n (%) | 2,752 (1.3) | 939 (1.3) | 938 (1.3) |
| Asian or Asian British, n (%) | 3,564 (1.7) | 1,266 (1.8) | 1,276 (1.8) |
| Chinese, n (%) | 606 (0.3) | 209 (0.3) | 207 (0.3) |
| Ever Smoke, n (%) | 96,277 (45.5) | 32,039 (45.4) | 32,171 (45.6) |
| FEV1 / FVC, mean (SD) | 0.76 (0.06) | 0.76 (0.06) | 0.76 (0.06) |
| Death Events, n (%) | 7,376 (3.5) | 2,536 (3.6) | 2,478 (3.5) |
| Time to Event, yr, mean (SD) | 11.2 (1.5) | 11.2 (1.5) | 11.2 (1.5) |

**Table 2:**
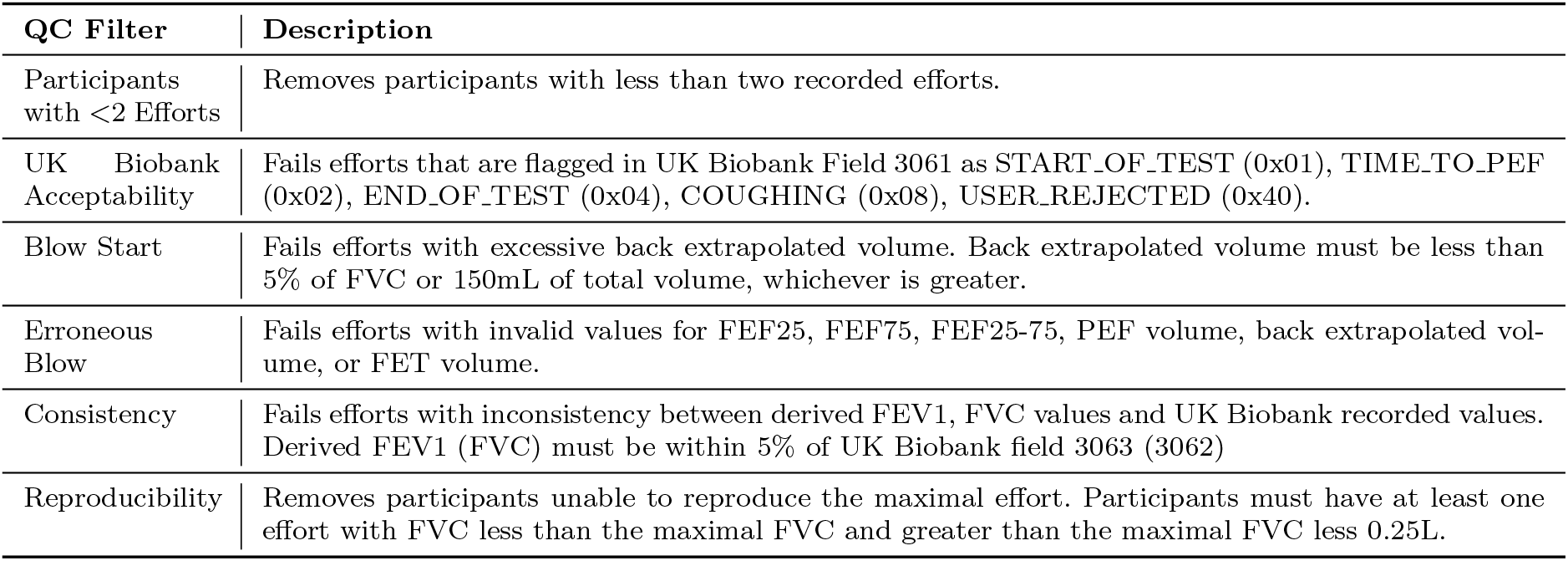
QC filter description for preprocessing participants and spirometry efforts from the UK Biobank.

| QC Filter | Description |
| --- | --- |
| Participants with <2 Efforts | Removes participants with less than two recorded efforts. |
| UK Biobank Acceptability | Fails efforts that are flagged in UK Biobank Field 3061 as START_OF_TEST (0x01), TIME_TO_PEF (0x02), END_OF_TEST (0x04), COUGHING (0x08), USER_REJECTED (0x40). |
| Blow Start | Fails efforts with excessive back extrapolated volume. Back extrapolated volume must be less than 5% of FVC or 150mL of total volume, whichever is greater. |
| Erroneous Blow | Fails efforts with invalid values for FEF25, FEF75, FEF25-75, PEF volume, back extrapolated volume, or FET volume. |
| Consistency | Fails efforts with inconsistency between derived FEV1, FVC values and UK Biobank recorded values. Derived FEV1 (FVC) must be within 5% of UK Biobank field 3063 (3062) |
| Reproducibility | Removes participants unable to reproduce the maximal effort. Participants must have at least one effort with FVC less than the maximal FVC and greater than the maximal FVC less 0.25L. |

**Fig. 1:**
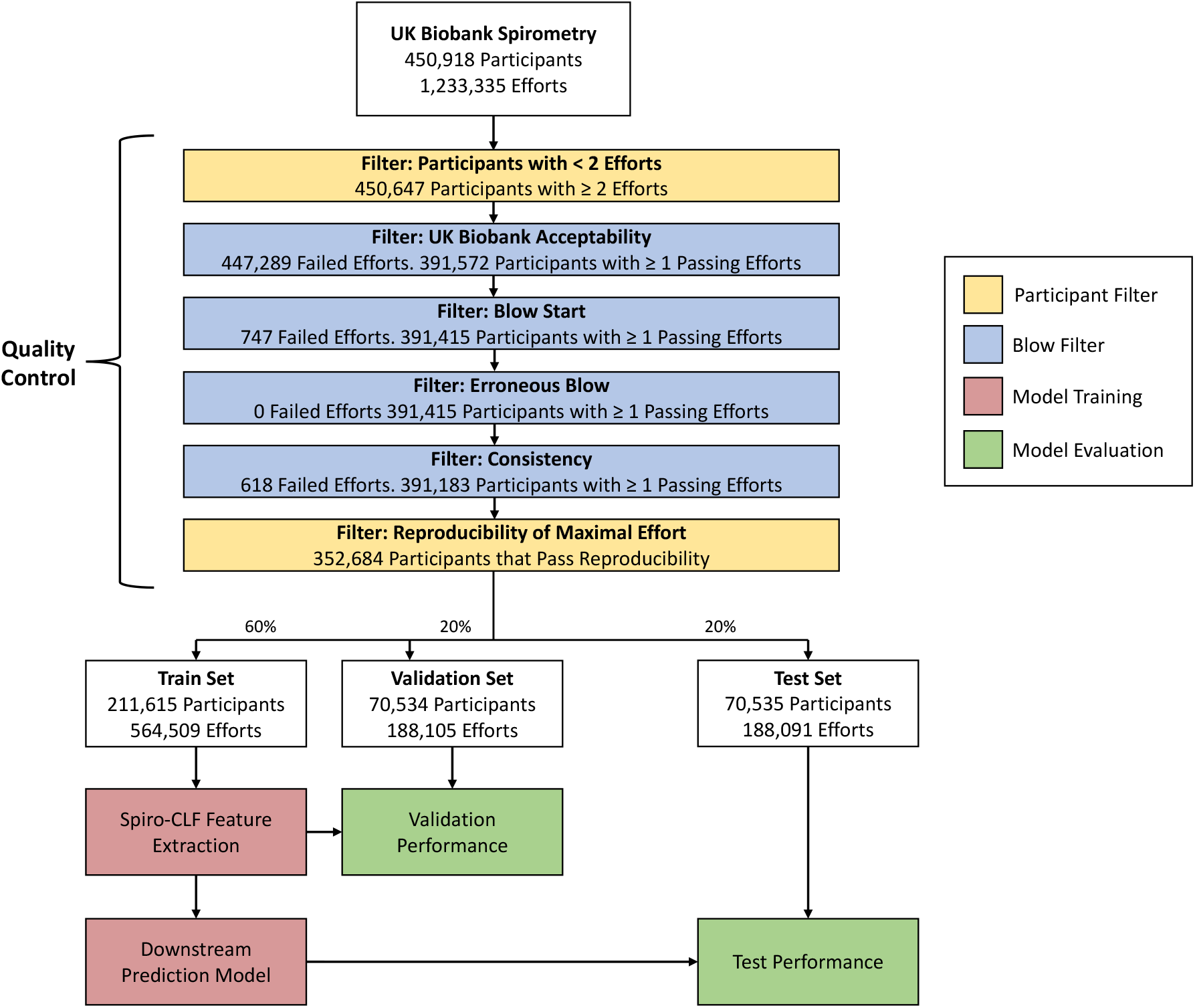
Overview of the Quality Control (QC), Spiro-CLF training, and downstream prediction process. The QC process contains participant filters, which contains participant-level criteria, and blow filters, which contain effort-level criteria. In each blow filter, spirometry efforts are tested against the specified criteria and labeled as either passing or failing the given filter. Participants with less than one QC-passing efforts or failing to produce a reproducible effort are removed from the dataset. QC criteria details are provided in Table 2.

After removing the participants with less than one QC-passing effort or fail the reproducibility criteria, the entirety of the efforts from remaining participants are included in the dataset. The distribution of remaining efforts is shown in Fig. 2.

**Fig. 2:**
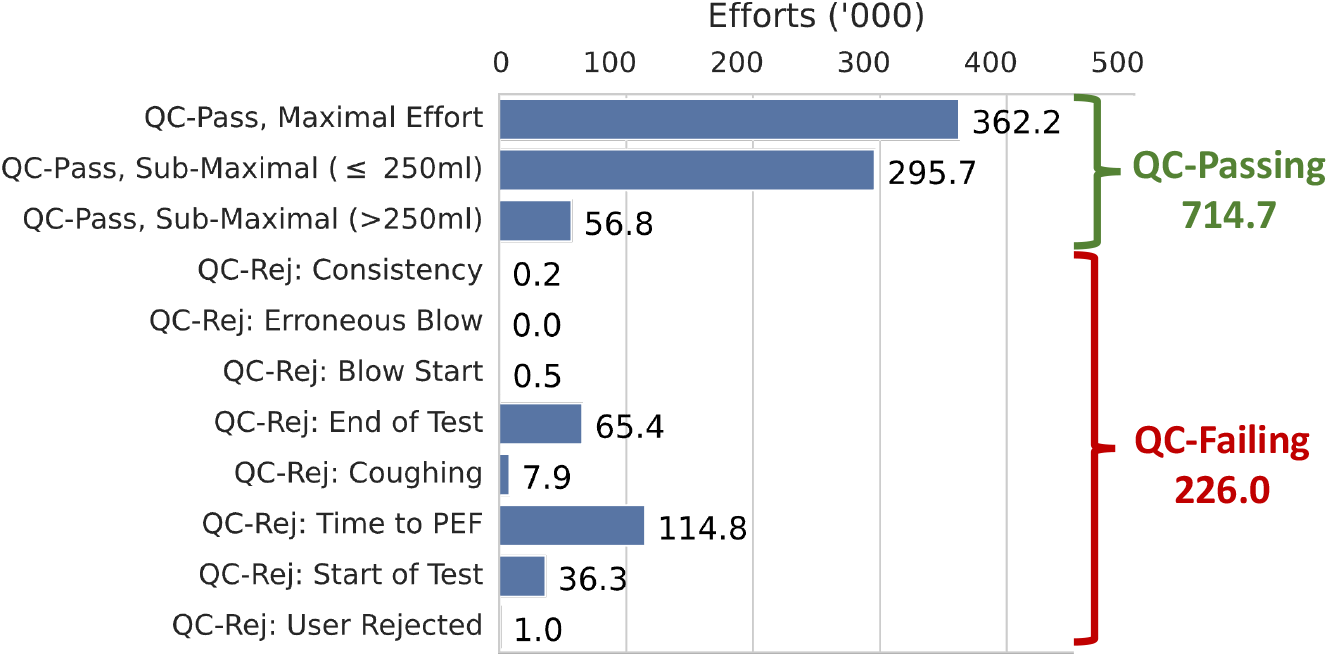
Number of efforts in the dataset after preprocessing, listed by effort type. *QC-Pass, Submaximal* ≤ *250mL* denotes efforts that pass QC and have FVC within 250mL of the maximal FVC.

After the QC process, the raw spirometry data was downsampled from 10ms to 50ms intervals and the exhalation duration was limited to 15s to improve model efficiency. Efforts were truncated after reaching FVC; we replaced the spirometry volume values after achieving FVC to be the derived FVC value for the effort.

### 2.2 Representation Learning with Spiro-CLF

The aim of our study was to uncover the complex, non-linear relationships between discarded spirometry data and the derived spirometry metrics, i.e. FEV_1_ and FVC, for UK Biobank participants. The raw spirometry efforts are often “noisy”, in that the observed efforts can be characterized as noisy signals from some underlying lung function. We therefore introduced a contrastive learning approach, **Spiro**gram-based **C**ontrastive **L**earning **F**ramework (Spiro-CLF) to identify a feature representation that is invariant to noise in each individual effort, thereby capturing the underlying factors that contribute to lung function.

Contrastive learning is a self-supervised approach that has recently shown to be effective in improving image and language processing prediction tasks (Jaiswal et al., 2020; Shurrab and Duwairi, 2022). It typically involves two steps: 1) an initial prediction task, often called the *pretext* task, that learns a feature representation of the data, and 2) the *downstream* task, which uses the learned representation on the ultimate prediction goal. The pretext task is self-supervised in that the model is discriminative, i.e. it is trained to predict a given label, however the pretext labels are derived entirely from the dataset itself.

Our work is most closely related to Chen et al. (2020), which was introduced in the context of image classification, and uses different correlated transformations of a given image as input to a deep learning model. Spiro-CLF defines each data sample as an individual participant, and each spirometry effort as different perspectives of that participant’s lung function. We additionally applied transformations (e.g. flow-volume, flow-time) to the spirometry efforts to further improve model performance; we define these transformations in section 2.4.

An overview of the Spiro-CLF representation learning process is shown in Figure 3. During training, Spiro-CLF sampled randomized batches of spirometry efforts. Each batch contained pairs of spirometry efforts produced from the same individual. Each effort within the batch had one positive example belonging to the same individual, and *B* − 2 negative examples, where *B* is the batch size. The model predicted whether any two given efforts within the batch were produced by the same individual or different individuals, represented by a predicted similarity score for each pair of efforts. The contrastive loss encouraged the model to predict high similarity for efforts produced from the same individual, and low similarity for efforts from disparate individuals.

**Fig. 3:**
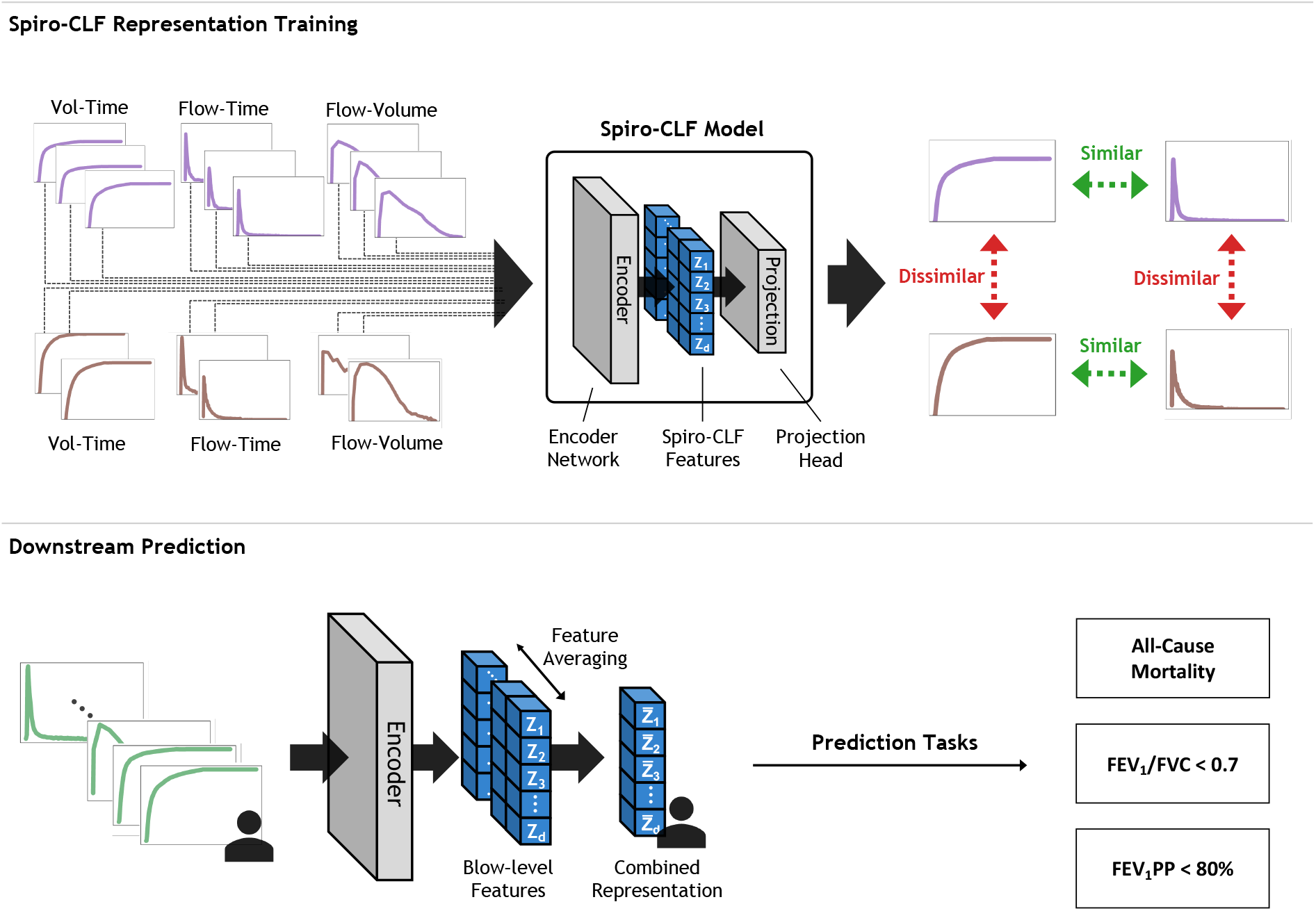
**Top:** Schematic of the Spiro-CLF training process. We randomly applied flow-time, flow-volume, and identity (volume-time) transformations to each spirometry effort within the training batch. The Spiro-CLF model is trained with contrastive loss to predict high pairwise similarity between efforts from the same individual and low pairwise similarity between efforts from different individuals. **Bottom:** Once the Spiro-CLF model is trained, we applied the encoder network to the entirety of an individual’s efforts, including transformation, to generate a combined feature representation. This representation can then be used in a variety of downstream predictive tasks.

We parameterized the Spiro-CLF model using a Convolution Neural Network (CNN) architecture, which captures the temporal relationship between each time step in each sample. CNN models have been previously shown to be highly effective on time-series prediction tasks (Hoseinzade and Haratizadeh, 2019; Wang et al., 2019). The convolution layers within CNNs encode the assumption that the relationship between values at adjacent time steps are more relevant than values between non-adjacent time steps. The model architecture and training process is defined in more detail in section 2.3.

Once the model is trained, we used the output of an intermediate layer as the learned representation of a participant’s overall lung function. We incorporated a feature averaging step (Foster et al., 2021) when generating Spiro-CLF features to further encourage invariance of the learned representations. During this step, we sampled the entirety of transformed efforts from each individual and averaged the respective features. The final representation can then be used in downstream prediction tasks; in particular we evaluated 1) binary prediction for FEV_1_/FVC < 0.7 and FEV_1_ Percent Predicted (FEV_1_PP) (Sec. 2.6), and 2) Cox regression for all-cause mortality (Sec. 3.3).

We used a bootstrap resampling procedure to estimate the standard deviation of the reported prediction performance. Both the Spiro-CLF model and downstream models were retrained on 24 bootstrapped training distributions and evaluated on the out-of-bag samples. The 95% confidence intervals for the performance scores are reported for each experiment.

### 2.3 Spiro-CLF Model

We parametrized the Spiro-CLF model with a deep neural network. Model architecture and hyperparameters were selected using a randomized grid search procedure, optimized with respect to minimum loss on the validation set. The model consisted of six convolution blocks; each block contained a 1-dimensional convolution layer with weight normalization, ReLU activation function, and skip-connection. Each convolution had a fixed kernel size of 40 with padding to avoid downsampling. After the convolution blocks, two fully-connected layers were applied to downsample to the 100-dimensional feature space. We trained an additional projection head parametrized by a multi-layer perceptron (MLP) with one hidden layer, ReLU activation, and batch normalization. The projection head maps the feature space to a 128-dimensional output to calculate contrastive loss. The detailed model architecture is shown in Appendix D Figure 10.

We trained the model for 400 epochs to minimize contrastive loss of the projection head output. During training, contrastive loss was calculated using batches of 512 data samples. For each batch, 256 unique participants were randomly selected, followed by the random selection of two transformed spirometry efforts from each selected participant. The available transformations are discussed in section 2.4. The model was applied to the samples, and a pairwise similarity score and contrastive loss was calculated between each of the 512, 100-dimensional, vector outputs of the projection head. The model was trained using the Adam optimizer (Kingma and Ba, 2014) with an initial learning rate of 10^−2^ and a scale factor of 0.1 on validation loss plateau.

After training, the projection head was discarded, and the model was applied to the spirometry samples in the test set to calculate the Spiro-CLF features.

### 2.4 Sampling of Spirometry Efforts and Transformations

During Spiro-CLF model training, we randomly selected two efforts from each participant in a given batch of participants. The two efforts were sampled uniformly from the participant’s collective spirometry efforts. If a participant had exactly two efforts, then both efforts were used in the training batch. We then applied a random data transformation to each effort.

Data transformations are non-learned mappings applied to the data samples to increase information on the underlying lung function. Each applied transformation returns different perspectives of a single participant’s lung function. During the data transformation stage, we applied either a flow-time transformation, flow-volume transformation, or identity (volume-time) transformation, selected with uniform probability. These transformations are detailed below. Therefore, in each training batch, each participant had two separate spirometry efforts, which were represented by either volume-time, flow-time, or flow-volume curves.

#### Flow-Time

We applied a flow-time transformation to the original flow-time data by calculating the change in expiratory volume over each time interval. We similarly downsampled the flow-time curves to 50ms intervals.

#### Flow-Volume

We applied a flow-volume transformation to the original flow-time data. Volumes were sampled in 10mL fixed intervals. Due to the 10ms sampling interval in the original flow-time data, the exact flow at each 10mL volume interval was not always available; we used linear interpolation to estimate the flow at those missing intervals.

### 2.5 Time-to-Event Prediction

We used UK Biobank field 3060 to identify the date the recorded spirometry was performed and field 40000 for event time for all-cause mortality. The dataset contained 12,390 mortality events (3.5%) with a median time-to-event of 11 years.

We trained a Cox Proportional Hazards Regression model on the Spiro-CLF features extracted from the training set and evaluated the concordance index (c-index) on the Spiro-CLF test features. C-index is a goodness-of-fit measurement that evaluates the concordance between the labels and predicted outcomes. A higher c-index indicates a better model fit. We omitted any covariates in order to evaluate the predictive power of the Spiro-CLF features. Interestingly, incorporating feature averaging on the generated Spiro-CLF features reduced predictive performance for the Cox model. We therefore omitted this step in this predictive task (see Sec.4 for additional discussion).

We compared the results from using the Spiro-CLF representation with previously proposed spirometry metrics, such as FEV_1_, FVC, and the forced mid-expiratory flow between 25% and 75% of the FVC (FEF25-75). These alternative metrics were calculated on the maximal QC-passing effort for each individual. The set of metrics from the training dataset were then used as predictors in the Cox Regression model.

### 2.6 Lung Function Impairment Prediction

We defined lung function impairment using two metrics: FEV_1_/FVC and FEV_1_ percent of predicted (FEV_1_pp). We calculated FEV_1_pp using GLI-2012 reference values (Quanjer et al., 2012) calculated using the Spiref^1^ python package. We took sex, height, age at recruitment, and self-reported ethnicity values from fields 31, 12144, 21022, 21000. Self-reported ethnicities were mapped to Caucasian or Other to be consistent with the GLI-2012 classifications.

To predict lung function impairment we created binary labels for each individual based on a threshold for each metric. Specifically we defined FEV_1_/FVC < 0.7 and FEV_1_pp < 80%, the Global Initiative for Chronic Obstructive Lung Disease (GOLD) spirometric criteria for moderate-to-severe airflow limitation in COPD (Vogelmeier et al., 2017) as indicative of lung function impairment. Using these definitions we trained logistic regression models for each task using the Spiro-CLF features as input. The regression models were trained with no regularization and no additional covariates. Model performance was assessed using the Area under the Receiving Operating Characteristic (AUROC) curve on the test set.

### 2.7 Feature Space Analysis

We use a dimension-reduction method, Uniform Manifold Approximation and Projection (UMAP) (McInnes et al., 2018), to project the Spiro-CLF representations down to two dimensions for visualization purposes. We randomly sampled 2000 participants and calculated the Spiro-CLF feature embeddings for the entirety of their respective efforts, including flow-time and flow-volume transformations. We then applied UMAP to project the 100-dimensional embeddings to two dimensions.

We evaluated the level of invariance induced by the Spiro-CLF model by investigating the feature space. The goal of the Spiro-CLF model training is to create a representation of lung function that is invariant to the inherent noise in repeated exhalation effort. An invariant representation would have minimal variance even when the individual efforts from a participant are not identical. We evaluated the empirical performance by calculating the average Euclidean distance in the feature space for each individual’s efforts. We first calculated the average Spiro-CLF representation for each individual, then averaged the Euclidean distance between each individual representation and the average representation.

### 2.8 Feature Importance Approximation

In order to understand how the Spiro-CLF model and subsequent downstream prediction tasks operate on the spirometry inputs, we applied feature importance methods to understand the relative importance of each individual timestamp of the raw spirometry input. We first combined the Spiro-CLF model and linear Lung Function impairment model (Section 2.6) into a single prediction pipeline. We then applied the Asymmetric Shapley Value (ASV) (Frye et al., 2020) method to calculate the feature importance values.

ASV has a number of properties that are suited for this application:

- *Post-hoc*. ASV can be applied to any prediction model after the model has been trained with no modification to the training process or model architecture.
- *Local*. ASV can be calculated for individual data samples. Traditionally, most feature importance methods are *global*, where they give a single explanation for the entire dataset and model. Using a local method allows the investigation of different subgroups within the data distribution by averaging over the relevant samples. For example, we can calculate ASV separately for flow-time, flow-volume, and volume-time transformations.
- *Asymmetric*. Due to the time-dependency in the spirometry efforts, the individual features within each sample contains asymmetric interactions. For example, in our dataset each feature represents a 50ms timestamp. Within each pair of timestamps, one timestamp occurs before the other. Therefore the relative importance of the latter timestamp is conditional on the former timestamp. We can enforce this relationship using ASV using a directed graph which we define prior to use.

We approximated ASV using a Monte Carlo sampling algorithm (Strumbelj and Kononenko, 2014), using 2000 permutations per data sample. We applied this approximation on 50 randomly sampled spirometry efforts for each of flow-time, flow-volume, and volume-time transformations. We then averaged over each transformation to obtain the subgroup-level explanation.

## 3 Results

All experiments were performed on a computing cluster using AMD EPYC 7302 16-Core processors and NVIDIA A100 GPUs. All source code is available at https://github.com/davinhill/Spiro-CLF.

### 3.1 Participant Characteristics

We included 352,684 participants in this study, with between 2-3 spirometry efforts per participant (66.7% with 3 efforts) resulting in 940,705 total efforts represented as volume-time curves. Participants were recruited across 23 assessment centers in the United Kingdom for the UK Biobank database. Additional details regarding the UK Biobank study were previously published in Sudlow et al. (2015). We utilized the spirometry records from each participant’s baseline assessment, which was collected between 2006 - 2010. A summary of participant characteristics is listed in Table 1.

### 3.2 Spiro-CLF Training

We randomly partitioned the participants in the dataset into training (80%), validation (20%), and test (20%) sets. Data transformations (Sec. 2.4) were applied during training and test stages, which effectively tripled the number of data samples from 940,669 efforts to 2,822,007. Model training continued until convergence of the validation loss.

We applied the unsupervised Spiro-CLF representation as a proxy for lung function on a variety of prediction models in place of traditional spirometry metrics. We evaluated the quality of the learned representation on two tasks. First, we evaluated the ability of the Spiro-CLF representation, derived from any given effort, to recover the maximal FEV_1_/FVC. Second, we evaluated the usefulness of the Spiro-CLF representation in predicting mortality as compared with other spirometry metrics.

### 3.3 Spiro-CLF Prediction of All-Cause Mortality

We used a mortality prediction task to validate that the Spiro-CLF representation contains additional relevant information for disease progression and health outcomes. We first trained a Cox time-to-event model on the Spiro-CLF representation and evaluated the model fit using the c-index. We then trained Cox regression models on alternative spirometry metrics to establish a baseline comparison. No additional covariates were included in the model training in order to compare the predictive power of each metric. Results are shown in Figure 4. The model trained on Spiro-CLF representations achieved a c-index of 0.654, which surpassed the performance of the second-highest performing single metric, FEV_1_/FVC, with c-index 0.599 (P ≤ 1.4 *×* 10^−37^). We additionally trained Cox regression models on combinations of alternative metrics. Using the combination FEV_1_, FVC, and FEV_1_/FVC increased c-index to 0.615. Including all competing metrics in the Cox model further improved c-index to 0.627, which was still exceeded in performance by the Spiro-CLF features (P ≤ 8.3 *×* 10^−23^).

**Fig. 4:**
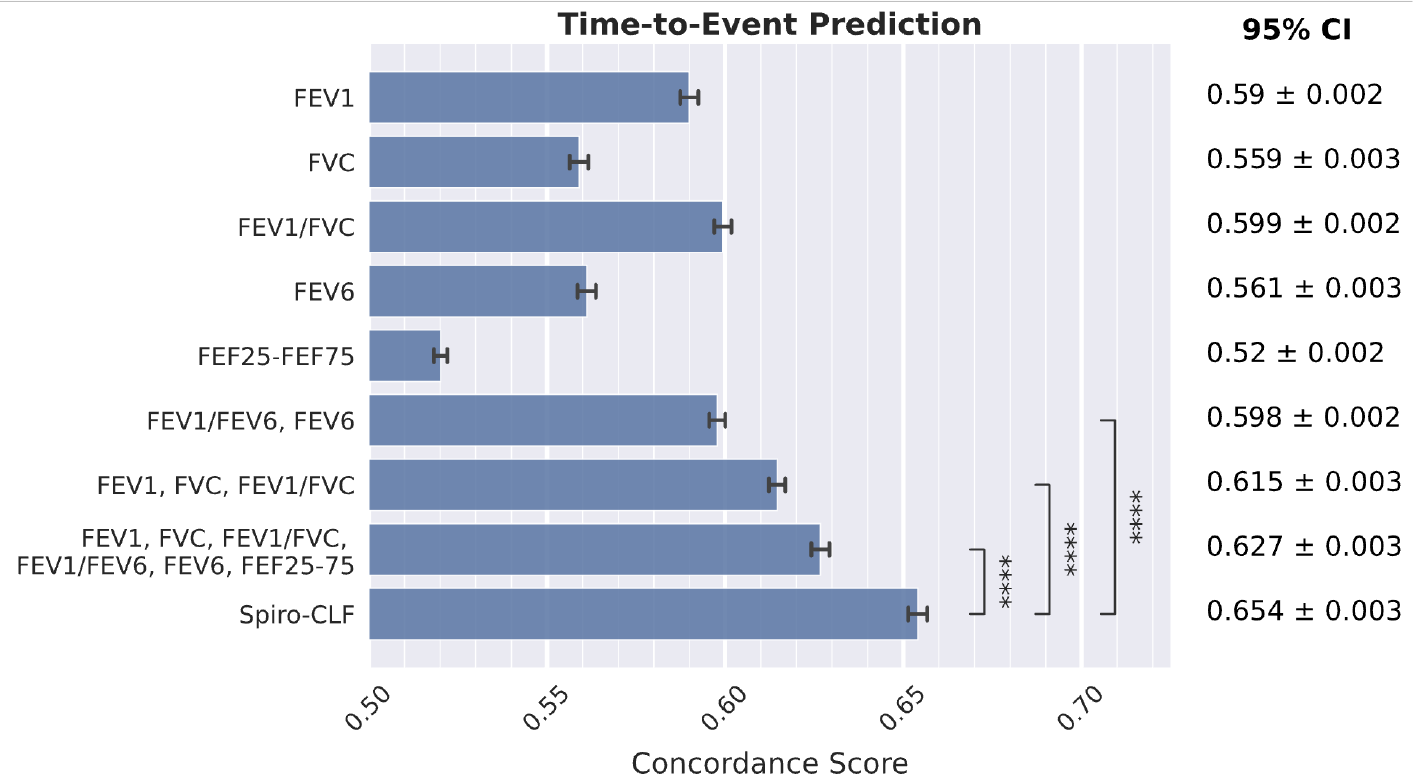
Results of a time-to-event mortality prediction task, measured using c-index (higher is better). We trained the Cox regression model using different lung function representations to compare their predictive power with respect to mortality prediction. The error bars represent 95% bootstrap confidence intervals (24 iterations). **** indicates significance at the Bonferroni-corrected 10^−4^ significance level.

### 3.4 Spiro-CLF Prediction of Lung Function Impairment

We compared the predictive power of the maximal, QC-passing effort while restricting the model to exclusively use submaximal and QC-rejected efforts. This comparison was evaluated on two lung function impairment prediction tasks: A) FEV_1_/FVC< 0.7 and B) FEV_1_PP< 80%. Results are shown in Figure 5.

**Fig. 5:**
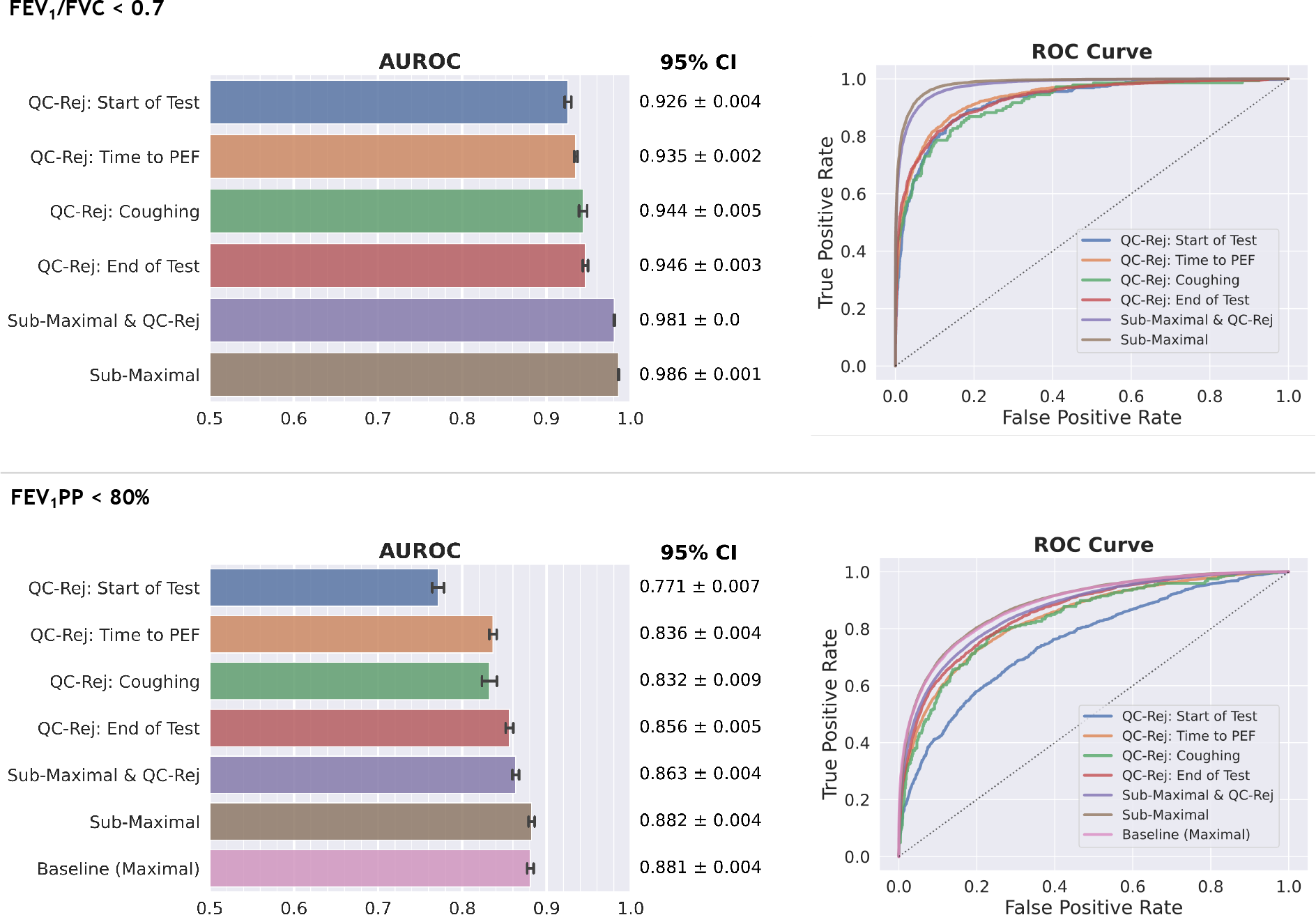
Results of the lung function impairment prediction tasks: FEV_1_/FVC< 0.7 (Top), and FEV_1_PP< 80% (Bottom). The error bars represent 95% bootstrap confidence intervals (24 iterations).

We observed that the Spiro-CLF model is able to achieve similar performance when using submaximal and QC-rejected efforts to the baseline on both the FEV_1_/FVC and FEV_1_PP prediction tasks.

We observed that the predictive performance of the Spiro-CLF model on QC-rejected and submaximal efforts was significantly recovered as compared to the baseline of using the maximal effort on both the FEV_1_/FVC and FEV_1_PP prediction tasks. In the FEV_1_/FVC task, the Spiro-CLF representations for submaximal and QC-rejected efforts achieved 0.981 AUROC. Exclusively using submaximal efforts yielded AUROC of 0.986. Similarly, in the FEV_1_PP prediction task we compare the performance of the submaximal and QC-rejected features to a baseline performance of using only features from maximal efforts. Using the Spiro-CLF features acheived 0.863 (submaximal and QC-rejected efforts) and 0.882 (submaximal efforts) AUROC compared to the baseline of 0.881 AUROC.

We additionally evaluated the performance when using only efforts that were QC-rejected for a variety of reasons. Restricting the Spiro-CLF model to using only efforts rejected due to excessive time to peak expiratory flow (PEF) recovered an AUROC of 0.935 for task A and 0.863 for task B.

### 3.5 Understanding the Spiro-CLF Feature Space

We visualized the feature embeddings generated by the Spiro-CLF model using dimension reduction methods. Given that the generated features are high-dimensional (100 dimensional vectors), we applied the Uniform Manifold Approximation and Projection (UMAP) (McInnes et al., 2018) algorithm on the output of the Spiro-CLF model. The resulting feature space visualization is shown in Figure 6.

**Fig. 6:**
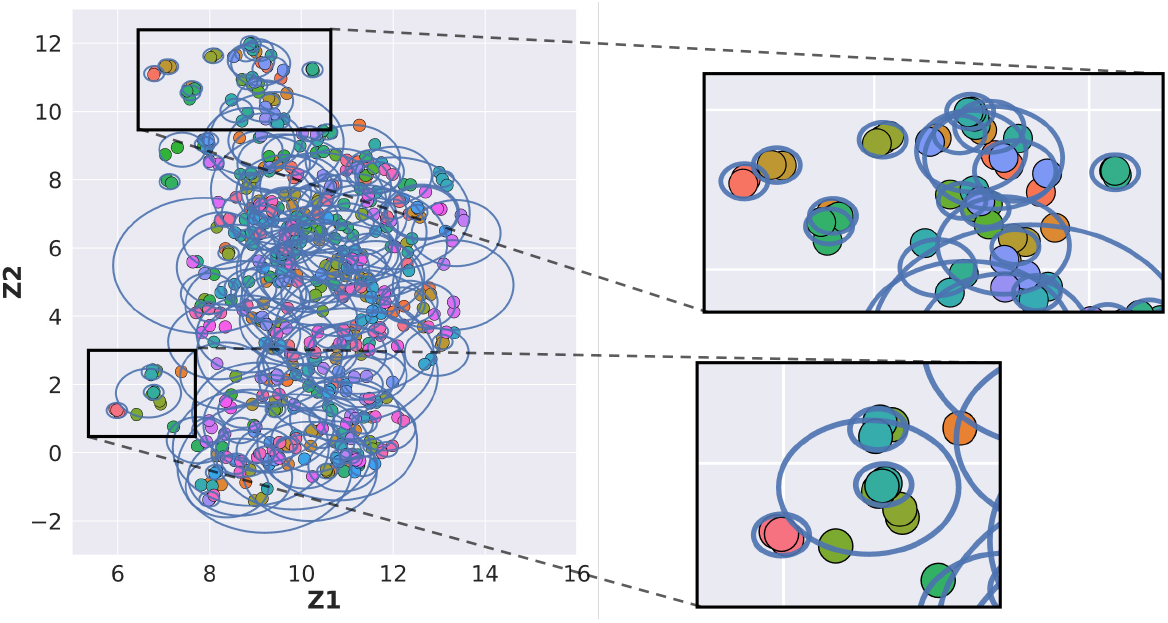
Analysis of the Spiro-CLF feature space which shows that efforts from the same individual are mapped to be close together in the feature space. We projected the Spiro-CLF representations of a random subset of 500 individuals to two dimensions using the UMAP algorithm. Each individual’s spirometry efforts were coded to be a different color and circled in blue.

The Spiro-CLF model was trained using a contrastive loss, which pushes spirometry efforts from the same individual closer together in the feature space. In the low dimensional feature space, Spiro-CLF spirometry representations resulting from different efforts or different data transformation appeared to project to a small locality. This observation indicates that the Spiro-CLF model generated similar feature embeddings for different spirometry efforts (or different transformations) performed by the same individual. We evaluated this observation by measuring the average Euclidean distance in the feature space generated using our trained model. Figure 6 is a visualization of the clusters of each individual efforts in the feature space. Each blue circle in the figure surrounds the features for a single individual. Table 3 calculates the distance between projections given different pairwise combinations of transformations. We observed that increasing the number of transformations increased the distance between clusters of feature projections. This indicates that adding transformations is a more difficult task for the Spiro-CLF model. Between the three investigated transformations, we observed that the flow-volume transformation yielded the largest average distance, which indicates that the model had the most difficulty in minimizing contrastive loss under this transformation.

**Table 3:**
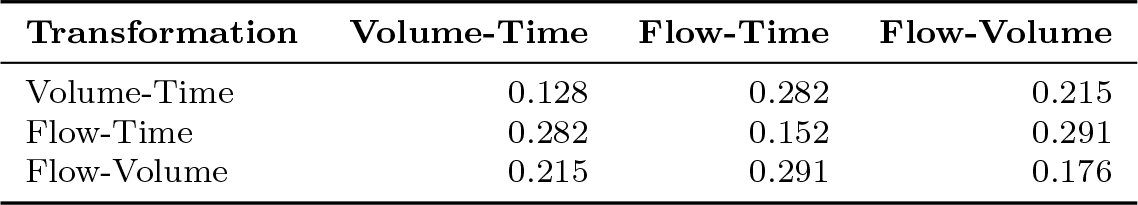
Effects of different transformations on the corresponding feature space. The table shows average distance between the feature representations of each individual’s efforts.

### 3.6 Assigning Relative Importance to Spirometry Curves

We used Asymmetric Shapley Values (ASV) to assign relative importance to the spirometry effort features for maximal efforts. We selected 50 randomized samples for each spirometry transformation (Fig. 7) and averaged ASV over each transformation. Importance measures are relative to the prediction of COPD in the downstream task. We observed that feature importance peaked within the first 1000 ms of the spirometry efforts, which is associated with peak expiratory flow (PEF) and the FEV_1_. This section of the importance curve corresponds to low variance in importance estimate over the averaged samples. In the flow-volume figure (Fig. 7(c)) we observed a decrease in importance between 2000mL and 4000mL segment with another local peak at 4100mL. This section corresponds to higher variance. This section generally corresponds to FVC, with higher variance due to sampled efforts achieving FVC at different timestamps.

**Fig. 7:**
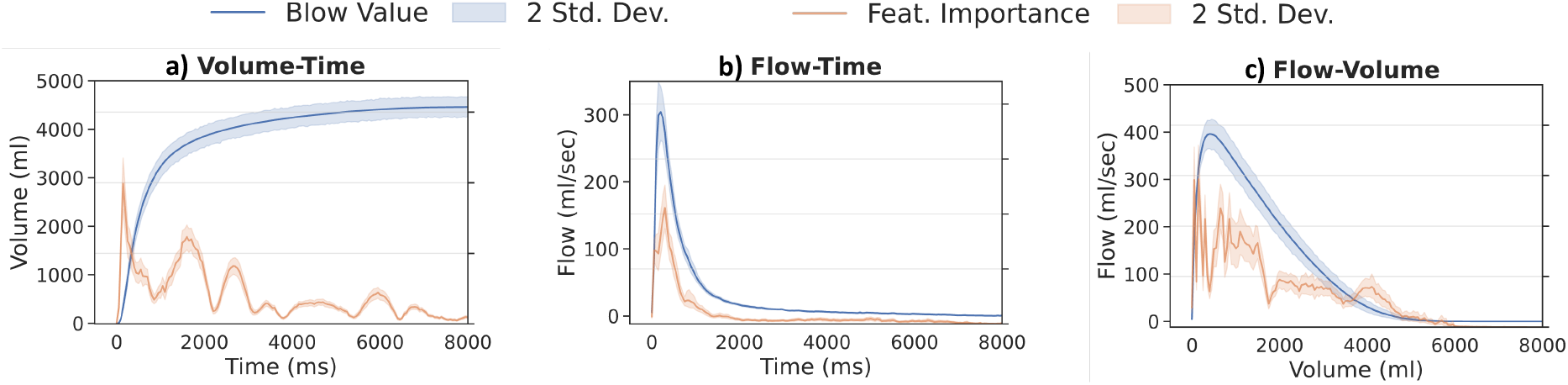
Visualization of the relative importance of different sections of the spirometry curve with respect to the prediction of lung function impairment.

## 4 Discussion

In this work, we trained a deep learning model on the raw spirometry curves in the UK Biobank study. We demonstrated that this Spiro-CLF model, trained on the entirety of each participant’s efforts, could not only predict lung function impairment and mortality when only using suboptimal or failed quality control data, but also improve prediction of mortality when added to maximal efforts.

Spiro-CLF features are a representation of lung function that are trained to be invariant to the associated noise within the spirometry test process. Therefore the Spiro-CLF features can be calculated for any effort from a given individual, including QC-rejected and submaximal efforts. Our model is able to maintain high prediction of lung function impairment using QC-rejected and submaximal efforts, and improve prediction of mortality when added to maximal efforts. In addition, the Spiro-CLF representation is generated from an unsupervised model which can be easily transferred across datasets. The resulting vector representation can be directly used as a replacement for traditional spirometry metrics in any prediction tasks. These data suggest that Spiro-CLF or other machine learning models could augment current clinical spirometry testing in providing more accurate prediction of airflow limitation, lung function, and other outcomes not only in optimal settings, but also when a sufficient number of efforts is unable to be obtained.

The Spiro-CLF model maps complex high-dimensional data to a rich lower-dimensional feature space. Existing literature in the machine learning field has shown that the learned Spiro-CLF feature space may encode other latent representations of the underlying data (Chen et al., 2021) and has been used for clustering (Li et al., 2021; Caron et al., 2020) as well as combined with generative models (Kim et al., 2021). Applying such methods to spirometry data could be a promising future direction for disease subtyping, disease progression, or combined with genome-wide association studies.

There are a number of possible causes for the improved predictive performance gained from using the Spiro-CLF representation. First, the Spiro-CLF encoding allows access to information from QC-rejected efforts. For each individual with rejected efforts, the rejected efforts may indicate underlying lung function impairment. For instance, elderly patients or those with severe lung disease may have difficulty performing a traditional FVC measurement appropriately and thus their spirometry efforts may more often be rejected (Vandevoorde et al., 2008; Bellia et al., 2008; Jing et al., 2009; Allen et al., 2010). For efforts that are not rejected, the additional submaximal efforts provide information related to the relative variance for the maximal effort. Encoding these sources of information can provide additional information not contained within the maximal effort.

In addition, the neural network parametrization of the Spiro-CLF model encodes nonlinear effects from the raw spirometry efforts. Previous works have shown that using neural networks, especially in conjunction with convolution layers, can improve representations of lung function with respect to prediction of COPD (Bhattacharjee et al., 2022), COPD subtypes (Bodduluri et al., 2020), and gene association (Cosentino et al., 2023; Yun et al., 2023).

Our models were trained on a large set of systematically obtained flow-volume curves available through UK Biobank. A significant factor in the predictive performance improvement likely resulted from the increase in training samples from utilizing the entirety of an individual’s efforts and additionally applying data transformations to obtain flow-time, flow-volume, and volume-time views of each effort. At a basic level, the combined efforts and transformations increase the number of training samples per individual, up to 9x more when compared with using the maximal effort and a single blow representation. Deep learning models are known to require a significant number of data samples during training (Vapnik, 1999) and increasing the number of samples and the variability in data views offered by volume-time transformations to flow-volume and flow-time enables the use of more complex models and reduces the likelihood of over fitting the training data.

We incorporated a feature averaging step for the Spiro-CLF features (Sec. 2.2), which improved performance for the FEV_1_/FVC< 0.7 and the Time-to-Event prediction tasks. However, using the averaged features for the FEV_1_PP< 80% reduced performance. The FEV_1_PPprediction label contains demographic information which is not available to the Spiro-CLF model, resulting in a more difficult prediction task. Since the Spiro-CLF model excludes covariate information, the resulting feature space does not disentangle these additional factors, which may result in worse performance when using feature averaging for tasks that depend on these covariates. For these tasks, the user should therefore omit the feature averaging step or include additional covariate information in the downstream prediction step.

Despite the increase of training samples used in the Spiro-CLF model, we acknowledge that overfitting is still possible, and we do not know if our model is transferable to clinical spirometry or other cohorts. The spirometry efforts in UK Biobank were limited to 3 efforts per individual and the QC definitions may be different from those used in a clinical setting. In order to compare our models to ‘ground truth’, we excluded participants that were unable to perform at least one QC-passing spirometry effort. The UK Biobank was predominantly of self-identified white race, European genetic ancestry, and - as a volunteer cohort - has a “healthy volunteer” selection bias (Fry et al., 2017). We did not evaluate the performance of our model for specific subsets of participants (age, race, chronic illness). Further validation in other cohorts and in clinical settings is needed.

Thus, for these reasons, additional training may be needed when applying the Spiro-CLF model to clinical spirometry. During training, additional validation may also be needed to ensure that model parameter and architecture choices are optimal with respect to the new dataset.

## Data Availability

This research has been conducted using the UK Biobank Resource under application number 20915. Feature representations and models produced in the present study are available upon reasonable request to the authors.

https://github.com/davinhill/Spiro-CLF

## A Acknowledgements

This research has been conducted using the UK Biobank Resource under application number 20915.

## B Funding

MHC is supported by NIH R01HL137927, R01HL135142, HL147148, and HL089856.

BDH is supported by NIH K08HL136928, U01 HL089856, and an Alpha-1 Foundation Research Grant.

DH is supported by NIH 2T32HL007427-41

PJC is supported by NIH R01HL124233 and R01HL147326.

SPB is supported by NIH R01HL151421 and UH3HL155806.

TY, FH, and CYM are employees of Google LLC

## C Disclosures

BDH receives grant support from Bayer.

MHC has received grant support from GlaxoSmithKline and Bayer, consulting fees from Genentech and AstraZeneca, and speaking fees from Illumina.

EKS has received grant support from GlaxoSmithKline and Bayer.

PJC has received grant support from Bayer.

SPB has received consulting fees from Sanofi/Regeneron and Boehringer Ingelheim, and CME fees from IntegrityCE. His institute has received funds from Sanofi and Nuvaira for the conduct of clinical trials.

TY, FH, and CYM are employees of Google LLC and own Alphabet stock.

## D Supplemental Figures

### D.1 Comparison of FEV_1_/FVC and FEV_1_PP prediction using the second-highest effort

In Section 3.4 we tested the effectiveness of using the Spiro-CLF representation for the binary prediction of A) FEV_1_/FVC < 0.7 and B) FEV_1_PP < 80%. However, the majority of QC-passing, submaximal efforts are within 250mL FVC of the maximal effort (Fig. 2). In Figure 8 we evaluated the effectiveness of using the second-highest effort (measured by FVC), in place of the maximal effort for the two binary prediction tasks. Specifically, we defined the ground-truth FEV_1_/FVC and FEV_1_PP values on the maximal effort and compared the agreement with the same values calculated on each individual’s second-highest, QC-passing, effort. The results are shown in a confusion matrix. We observe that using the second-highest effort leads to misclassification rates of 5.06% (0.832 F1) for task A and 5.23% (0.881 F1) for task B.

**Fig. 8:**
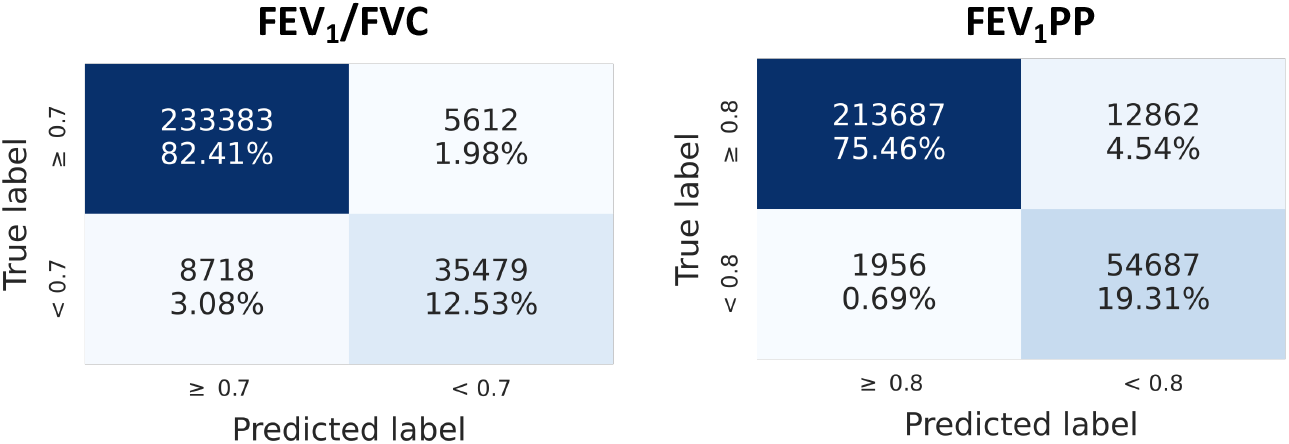
Confusion matrix when using the FEV_1_, FVC, and FEV_1_PP of the second-highest, QC-passing effort to predict FEV_1_/FVC < 0.7 and FEV_1_PP < 80%.

### D.2 Comparison of FEV_1_/FVC < 0.7 prediction using QC-rejected and submaximal efforts

In this experiment, we compared the results of the Spiro-CLF prediction of FEV_1_/FVC< 0.7 with the FEV_1_/FVCvalues of each subset of efforts. Specifically, we calculated the FEV_1_/FVCfor individual efforts in each QC-rejected and submaximal effort subset and used these values as predictions for the FEV_1_/FVC< 0.7 prediction task. We evaluate results using F1 score in place of AUROC due to the non-probabilistic property of the predictions. We observe in Figure 9 that using the Spiro-CLF representation improves prediction performance of all spirometry effort subsets (P ≤ 2.8 *×* 10^−6^).

**Fig. 9:**
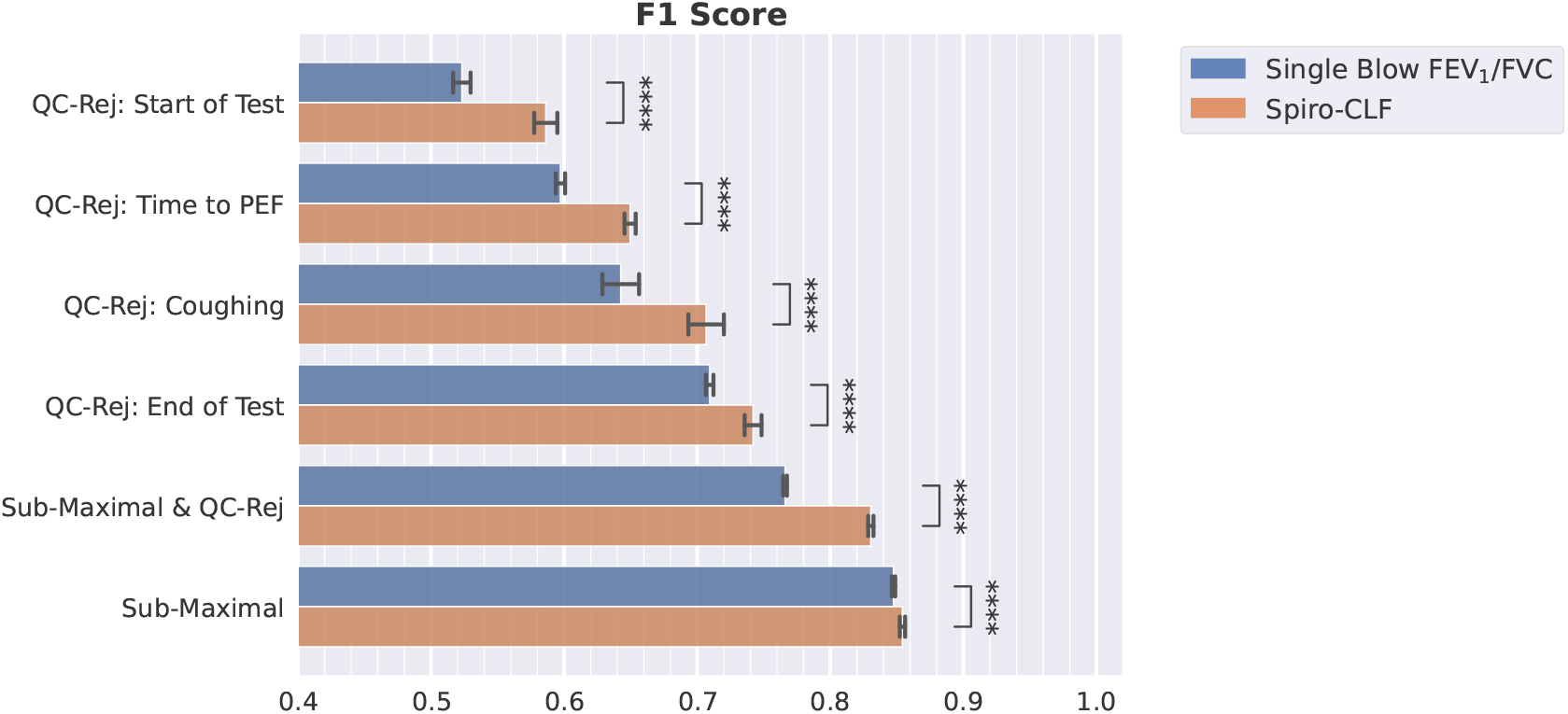
F1 score for FEV_1_/FVC < 0.7 prediction task. We compared the performance of the Spiro-CLF representation and the raw single-effort FEV_1_/FVC of each subset of efforts. The error bars represent 95% bootstrap confidence intervals (24 iterations). **** indicates significance at the Bonferroni-corrected 10^−4^ significance level.

**Fig. 10:**
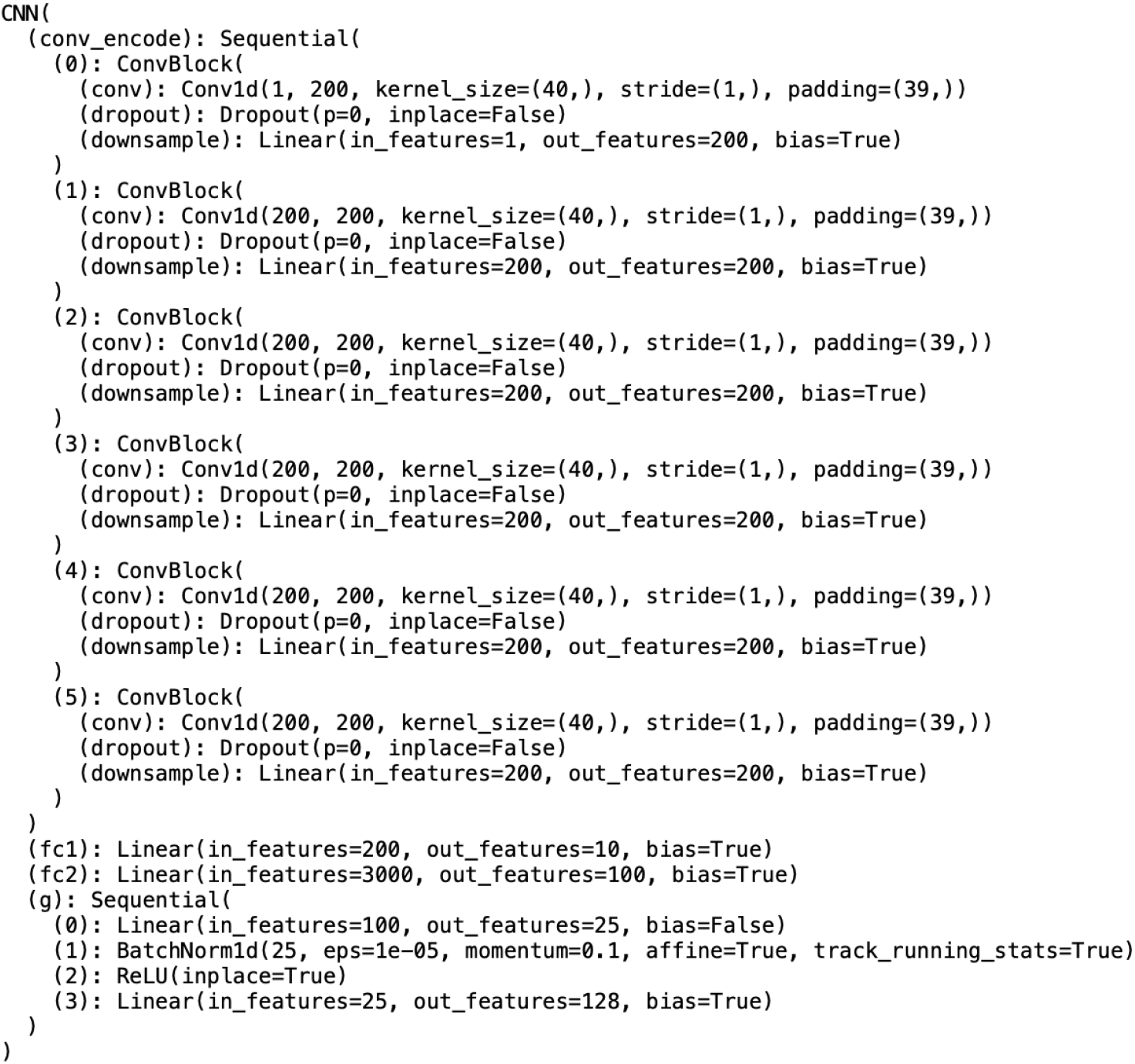
Model Architecture for Spiro-CLF model.

**Fig. 11:**
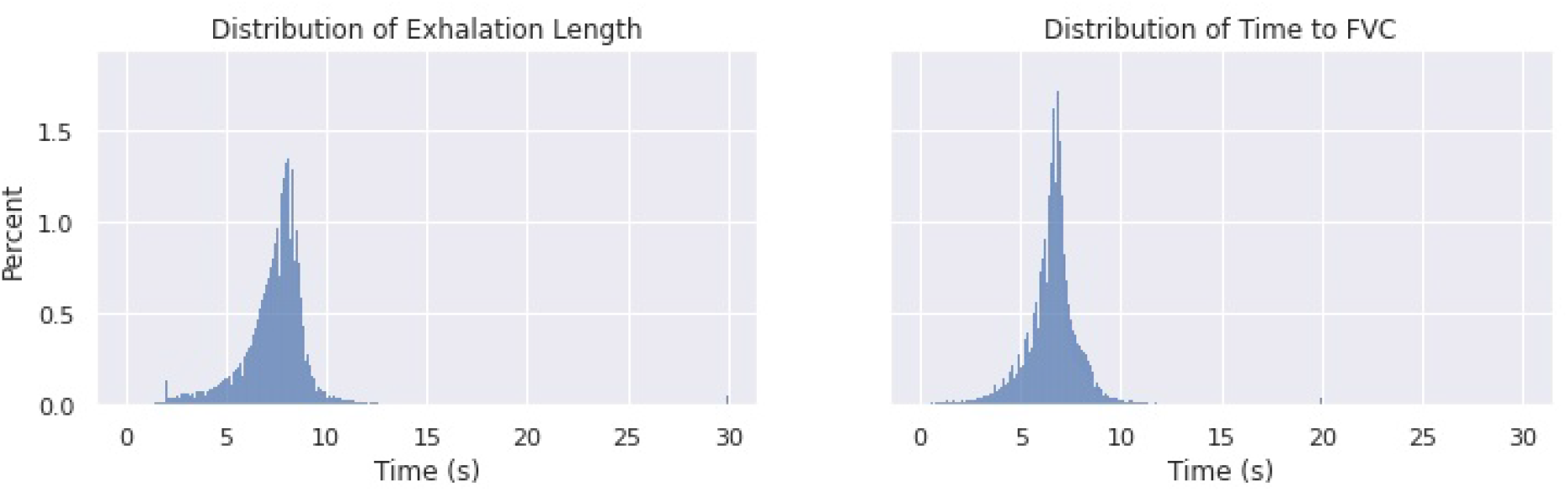
Distribution of exhalation length and time to FVC in the dataset.

## Footnotes

1 https://github.com/kennethverstraete/spiref

## Notes

### Author Declarations

This study used only publicly available data from the UK Biobank Resource.

